# The Natural Course of Peri-graft Fluid Collections After Kidney Transplantation: A Serial Imaging Cohort Study

**DOI:** 10.64898/2026.10.15.26364900

**Authors:** Malik Muhammad Makki, Komal Kumar, Harutyun Davtyan, Jiro Kusakabe, Eduardo Fernandes, Antonio Pinna, Khaled Refaai

## Abstract

**Background:** Fluid collections are common after kidney transplantation, but their prevalence at scheduled imaging and their management are inconsistent from canter to canter. We describe the cross-sectional prevalence and management of peri-graft collections, including an exploratory deep-drain subgroup.

**Methods:** In this retrospective single-canter cohort, 146 consecutive adult kidney transplant recipients (Cleveland Clinic Weston Hospital, July 2024 to June 2025) were imaged on postoperative day 0/1 and at approximately 1 and 3 months. Routine deep drainage was avoided (n=124); 22 clinically selected recipients with perioperative anticoagulation received a deep drain.

The primary outcome was cross-sectional prevalence of a peri-graft collection at each visit; secondary outcomes were management among patients with a collection. Drain-group comparisons were exploratory.

**Results:** Collections were present in 29 of 146 recipients at postoperative day 0/1 (19.9%; 95% CI, 14.2–27.1), 71 of 146 at 1 month (48.6%; 95% CI, 40.7–56.7), and 34 of 146 at 3 months (23.3%; 95% CI, 17.2–30.8). Prevalence in the no-drain group was 21.0%, 50.0%, and 23.4%, respectively; corresponding prevalence in the deep-drain group was 13.6%, 40.9%, and 22.7%.

Between-group prevalence comparisons were not statistically significant at any time point.

BMI was higher in the deep-drain group than in the no-drain group (30.04 vs 27.1 kg/m²; P=0.01). Among patients with a 1-month collection, management did not differ significantly between groups (any invasive management, 11.1% vs 16.1%; P=1.00). Among patients with a 3-month collection, invasive management was more common in the deep-drain group than in the no-drain group (80.0% vs 13.8%; P=0.007). This comparison included only five deep-drain recipients and was confounded by clinical selection, perioperative anticoagulation, and BMI imbalance.

**Conclusions:** Peri-graft collection prevalence was highest at 1 month and lower at 3 months in this predominantly no-drain cohort. Most collections were managed conservatively, and invasive treatment was uncommon in the no-drain group. The deep-drain subgroup provides clinically relevant information about high-risk recipients selected for drainage but cannot support causal conclusions regarding drain benefit or harm. Patient-level linkage is required to distinguish persistence, resolution, and new collection formation.

## Introduction

Peri-graft fluid collections are among the commonest surgical complications after kidney transplantation. They include lymphoceles, hematomas, urinomas, seromas and abscesses, and reported lymphocele incidence ranges from 0.6% to 51%^1,2^, largely because studies differ in definition, imaging schedule and follow-up. A consensus group found that only 32 of 87 published studies defined the outcome and none graded its severity^3^. Most collections are small and asymptomatic, but a minority obstruct the ureter, compress the iliac vein, impair graft function or become infected^4,5^, and lymphocele has been linked to hospital readmission^6^.

Closed-suction drains have long been placed to evacuate fluid and to give early warning of leak or bleeding, but their benefit is uncertain and they may offer a route for infection in immunosuppressed patients.^7^. Meanwhile, prevention strategies such as peritoneal fenestration, lymphatic ligation techniques and sclerosants alter collection rates^2,14,15^, which complicates interpretation of the natural history of collections.

A drain-free strategy therefore raises a practical question that the literature answers poorly: what happens to a collection when nothing is done to it? Most published series capture collections that were symptomatic, drained or treated. ^7^ Far less is known, at fixed early time points, about how often collections appear, when they peak, how many resolve without intervention and how many escalate to image-guided or operative treatment. Answering this would let surgeons set surveillance intervals and intervention thresholds and would reassure clinicians who default to intervening on small asymptomatic collections.

Perioperative anticoagulation adds a second unknown. It has been associated with a higher lymphocele rate^16^, and many surgeons place a deep drain in anticoagulated recipients. Whether that changes the trajectory of collections is unclear. We therefore aimed to describe the observed prevalence trajectory of peri-graft collections after kidney transplantation with serial imaging at postoperative day 0/1, 1 month and 3 months in patients managed without routine deep drainage and, secondarily and exploratorily, to compare this pattern with recipients who received a deep drain because of perioperative anticoagulation.

## Methods

### Study design and setting

This was a retrospective single-center cohort study at Cleveland Clinic Weston Hospital, Florida. The study was approved by the institutional review board and the requirement for individual consent was waived.

### Participants

All consecutive adult kidney transplant recipients transplanted between July 2024 and June 2025 were eligible. A total of 146 recipients were included and followed for 3 months. No additional exclusions were applied beyond consecutive eligibility (Figure 1).

**Figure 1.**
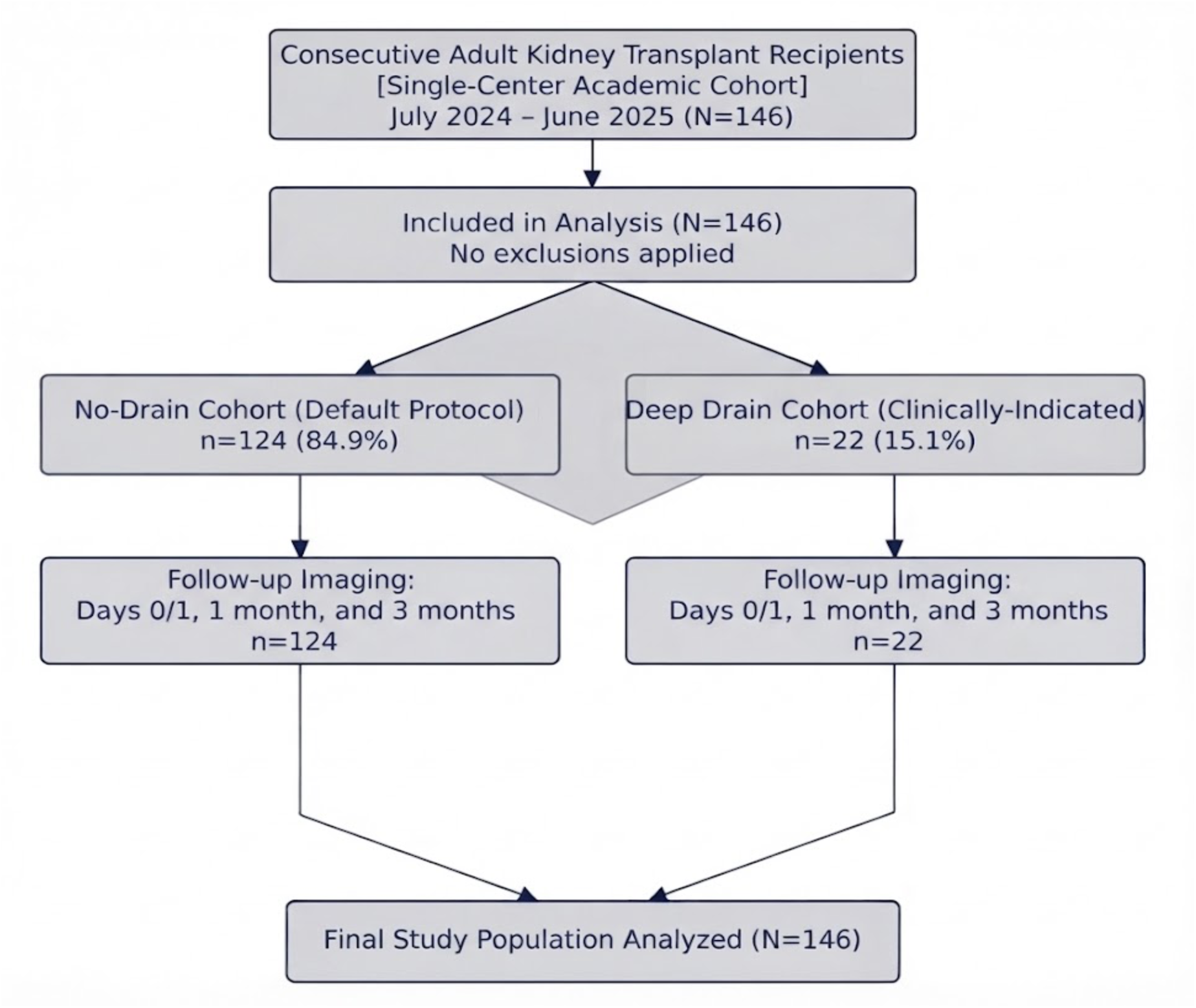
Study flow diagram. Consecutive adult kidney transplant recipients were managed with a predominantly no-drain strategy or received a deep drain selectively in the setting of perioperative anticoagulation.

### Drain strategy in Kidney Transplantation

The default strategy was avoidance of routine deep drainage. A no-drain protocol was the default (n=124). A deep drain was selectively placed in 22 recipients because of perioperative clinical risk, including perioperative anticoagulation. Drain assignment was not randomized and did not represent a treatment allocation. The deep-drain recipients were therefore considered an indication-enriched, clinically selected subgroup, not a comparison arm.

### Imaging surveillance and outcome definitions

A screening ultrasound was obtained on postoperative day 0/1, followed by ultrasound or CT at approximately 1 and 3 months. A peri-graft collection was defined as any fluid collection adjacent to the transplanted kidney on imaging.

Management was classified as conservative management, image-guided drainage, open surgical drainage, or any invasive management (image-guided drainage or open surgery combined). Collections without a documented image-guided or surgical procedure were reclassified as conservatively managed only after chart confirmation of the absence of a procedure, rather than being assumed conservative from absent documentation alone.

The study focused on peri-graft collections broadly, however, collection-specific type including lymphoceles, hematomas, urinomas, seromas and abscesses. and severity grading were not included in this analysis. Collection size, location and symptoms were extracted from radiology and clinic records by two reviewers, describe how disagreements were resolved.

### Outcomes

**Primary outcome** was the cross-sectional prevalence of a peri-graft collection at postoperative day 0/1, 1 month and 3 months.

**Secondary outcomes** were management category among patients with a collection at the 1-month and 3-month visits. Readmission, infection and other graft-related complications were not analysed in this study. Serum creatinine and estimated glomerular filtration rate were not assessed at the patient level, and no inference about renal function is drawn from this analysis.

### Bias and study size

Consecutive inclusion limited selection bias, and scheduled imaging reduced ascertainment bias relative to symptom-triggered series. Screening on day 0/1 detects small early fluid that may be clinically irrelevant, so day 0/1 findings are reported separately from the 1-and 3-month findings. Drain assignment was determined by anticoagulation and other perceived perioperative risk, which introduces confounding by indication in the drain comparison; the deep-drain group also had a significantly higher mean BMI (Table 1), a second axis of confounding. All eligible patients were included, and no sample-size calculation was performed; precision is reported through 95% confidence intervals.

**Table 1.** Baseline characteristics by drain status.

| Characteristic | Overall (n=146) | No drain (n=124) | Deep drain (n=22) | P |
| --- | --- | --- | --- | --- |
| Age, years, mean | 58.8 | 58.8 | 59.1 | 0.94 |
| Male sex, n (%) | 89 (61.0) | 75 (60.5) | 14 (63.6) | 0.88 |
| Race, n (%) |  |  |  | 0.76 |
| White | 46 (31.5) | 40 (32.3) | 6 (27.3) |  |
| Black | 63 (43.2) | 52 (41.9) | 11 (50.0) |  |
| Hispanic | 19 (13.0) | 16 (12.9) | 3 (13.6) |  |
| Asian | 14 (9.6) | 13 (10.5) | 1 (4.5) |  |
| Other / not recorded | 4 (2.7) | 3 (2.4) | 1 (4.5) |  |
| BMI, kg/m<sup>2</sup>, mean | 27.1 | 27.1 | 30.04 | 0.01 |
| Perioperative anticoagulation, n (%) | 22 (15.1) | 0 (0) | 22 (100) | — |

### Statistical analysis

Categorical variables were summarized as number and percentage. Continuous variables were summarized as means.The standardized mean difference for male sex was calculated from the reported group proportions. Baseline P values were obtained from the study database analysis (t test for means and chi-square test for race).

Overall prevalence is reported with Wilson 95% confidence intervals^18^. Drain-group comparisons of prevalence and management used Fisher’s exact test (two-sided). A two-sided P value below 0.05 was considered statistically significant, but all comparisons are exploratory, unadjusted, and not corrected for multiplicity.

The analysis was organized as follows. The primary analysis is collection prevalence in the entire cohort at each time point (Table 2). The secondary analysis is management among patients with a collection (Table 4). The comparison of the deep-drain and no-drain groups (Tables 3 and 4) is exploratory and does not support causal conclusions about drains, because drain assignment was determined by clinical risk. No single drain-group comparison was designated as a primary outcome.

**Table 2.** Overall prevalence of peri-graft collections (primary analysis)

| Time point | Collection, n/N (%) [95% CI] | No collection, n/N (%) [95% CI] |
| --- | --- | --- |
| Postoperative day 0/1 | 29/146 (19.9) [14.2–27.1] | 117/146 (80.1) [72.9–85.8] |
| 1 month | 71/146 (48.6) [40.7–56.7] | 75/146 (51.4) [43.3–59.3] |
| 3 months | 34/146 (23.3) [17.2–30.8] | 112/146 (76.7) [69.2–82.8] |

CI, Wilson 95% confidence interval; entire cohort (N=146). Collection prevalence is the primary outcome

**Table 3.** Collection prevalence by drain status (exploratory)

| Time point | No drain | Deep drain | Fisher P |
| --- | --- | --- | --- |
| Postoperative day 0/1 | 26/124 (21.0) | 3/22 (13.6) | 0.57 |
| 1 month | 62/124 (50.0) | 9/22 (40.9) | 0.49 |
| 3 months | 29/124 (23.4) | 5/22 (22.7) | 1.00 |

Exploratory comparison; drain assignment was clinically selected, not randomized.

**Table 4.** Management among patients with a peri-graft collection (secondary analysis)

| Category, n (%) | Overall | No drain | Deep drain | Fisher P |
| --- | --- | --- | --- | --- |
| **1-month collection** | **N=71** | **N=62** | **N=9** |  |
| Conservative | 60 (84.5) | 52 (83.9) | 8 (88.9) | 1.00 |
| Image-guided drainage | 8 (11.3) | 8 (12.9) | 0 (0) | 0.58 |
| Surgery (open/laparoscopic) | 3 (4.2) | 2 (3.2) | 1 (11.1) | 0.34 |
| All invasive management | 11 (15.5) | 10 (16.1) | 1 (11.1) | 1.00 |
| **3-month collection** | **N=34** | **N=29** | **N=5** |  |
| Conservative | 26 (76.5) | 25 (86.2) | 1 (20.0) | 0.007 |
| Image-guided drainage | 7 (20.6) | 4 (13.8) | 3 (60.0) | 0.048 |
| Surgery (open/laparoscopic) | 1 (2.9) | 0 (0) | 1 (20.0) | 0.15 |
| All invasive management | 8 (23.5) | 4 (13.8) | 4 (80.0) | 0.007 |

Percentages use patients with a collection at the relevant visit as the denominator. Collections without a documented image-guided or surgical procedure are classified as conservatively managed, Fisher's exact test, two-sided, unadjusted and not corrected for multiplicity so the P=0.007 and P=0.048 findings should be interpreted with that multiplicity in mind.

Management at 1 or 3 months was analysed only among patients with a collection at that time point, with patients, not collections, as the unit of analysis. Propensity-score weighting was not performed because the deep-drain group was small and the patient-level covariates required for balancing were unavailable.

## Results

### Cohort characteristics

Of 146 recipients, 124 (84.9%) were managed without a deep drain and 22 (15.1%) received a deep drain. The deep-drain subgroup was clinically selected because of perioperative anticoagulation and other perceived risk factors.

Mean age was similar in the no-drain and deep-drain groups (58.8 vs 59.1 years; P=0.94). Male sex was present in 75 no-drain recipients (60.5%) and 14 deep-drain recipients (63.6%; P=0.88). Race distribution did not differ significantly between groups (P=0.76). Mean BMI was significantly higher among deep-drain recipients than no-drain recipients (30.04 vs 27.1 kg/m²; P=0.01), a second axis of confounding by indication alongside anticoagulation itself (Table 1).

### Primary analysis: Cross-sectional Prevalence peri-graft collections

Collections were present in 29 of 146 recipients (19.9%; 95% CI, 14.2–27.1) on postoperative day 0/1. Prevalence increased to 71 of 146 (48.6%; 95% CI, 40.7–56.7) at 1 month and decreased to 34 of 146 (23.3%; 95% CI, 17.2–30.8) at 3 months (Table 2; Figure 2).

**Figure 2.**
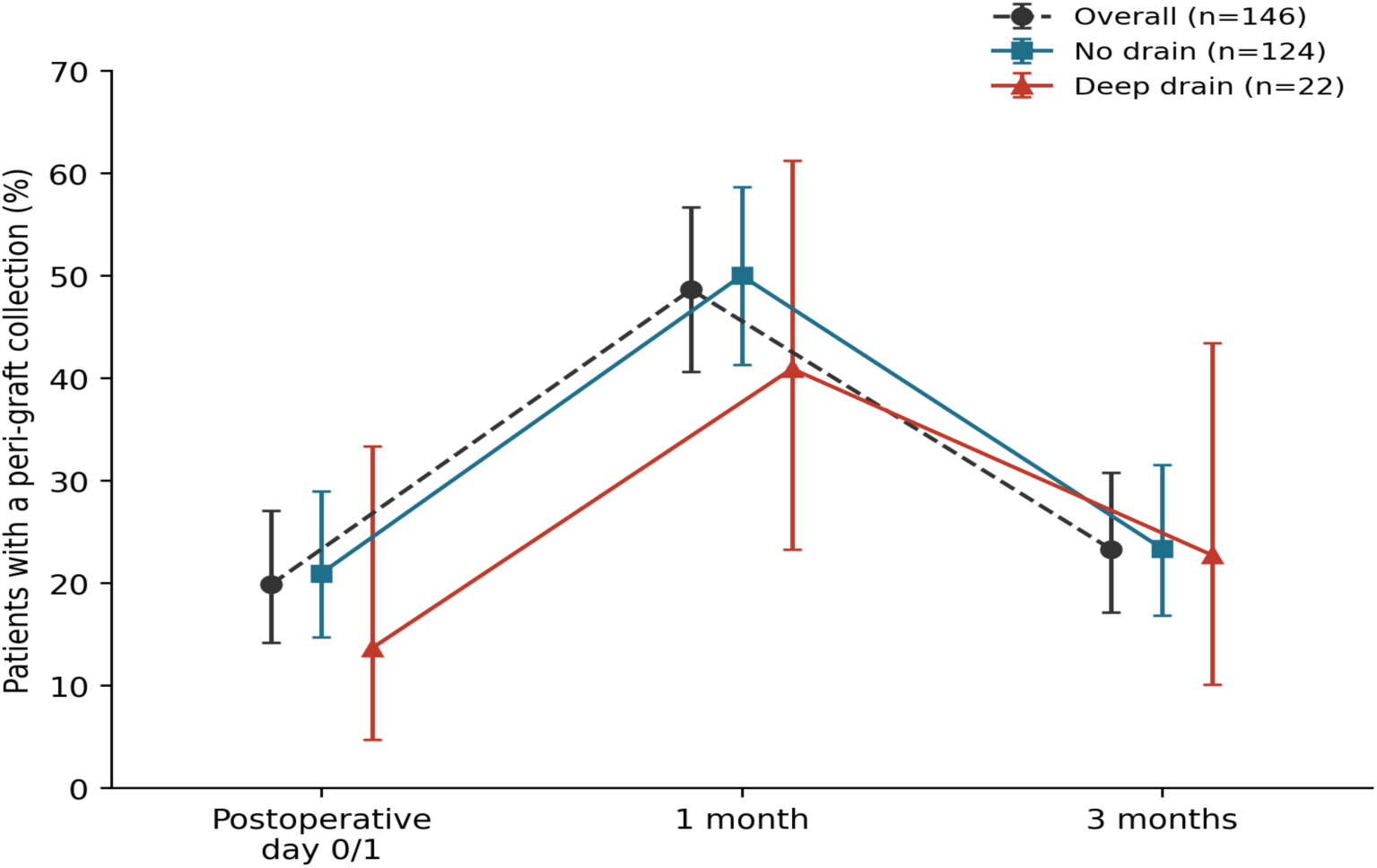
Cross-sectional prevalence of peri-graft collections on postoperative day 0/1, at 1 month and at 3 months, overall and by drain group. Points show observed prevalence and bars show Wilson 95% confidence intervals. All 146 recipients were imaged at each time point. Values are cross-sectional prevalence estimates and do not represent individual-patient incidence, persistence, or resolution.

### Summary of the Observed prevalence pattern, entire cohort

About one in five recipients (19.9%) has a collection on postoperative day 0/1; prevalence peaks at 1 month (48.6%) and falls to 23.3% by 3 months.

Given that, most 1-month collections are newly formed rather than persistent early collections, given that at least 42 of the 71 affected patients lacked a collection on day 0/1

The observed prevalence approximately doubled between postoperative day 0/1 and 1 month and then declined to a level near the initial postoperative prevalence by 3 months. These are cross-sectional estimates and should not be interpreted as individual-patient persistence or resolution.

### Exploratory prevalence comparison by drain status

In the no-drain group, collection prevalence was 21.0% at postoperative day 0/1, 50.0% at 1 month, and 23.4% at 3 months. In the deep-drain group, corresponding prevalence was 13.6%, 40.9%, and 22.7%. There was no statistically significant difference between groups at any time point (Table 3). With only 22 deep-drain recipients, these comparisons have limited power.

### Secondary analysis: management among patients with a collection

Management analyses used the number of patients with a collection at that visit as the denominator. Overall, at 1 month, 60 of 71 patients with a collection (84.5%) were managed conservatively, 8 (11.3%) underwent image-guided drainage, and 3 (4.2%) underwent open surgery. At 3 months, 26 of 34 patients with a collection (76.5%) were managed conservatively, 7 (20.6%) underwent image-guided drainage, and 1 (2.9%) underwent open surgery.

By subgroup, among no-drain patients with a 1-month collection, 52 of 62 (83.9%) were managed conservatively, 8 (12.9%) required image-guided drainage and 2 (3.2%) required surgery (10 of 62, 16.1%, required either); this did not differ from the deep-drain group (1 of 9, 11.1%; Fisher P=1.00). Among no-drain patients with a 3-month collection, 25 of 29 (86.2%) were managed conservatively and 4 of 29 (13.8%) required image-guided drainage, with no reoperations; among the 5 deep-drain patients with a 3-month collection, 4 (80.0%) required image-guided drainage or surgery, compared with 4 of 29 (13.8%) in the no-drain group (Fisher P=0.007). This comparison involves only 5 deep-drain patients selected by anticoagulation status and is exploratory (Table 4; Figure 3).

**Figure 3.**
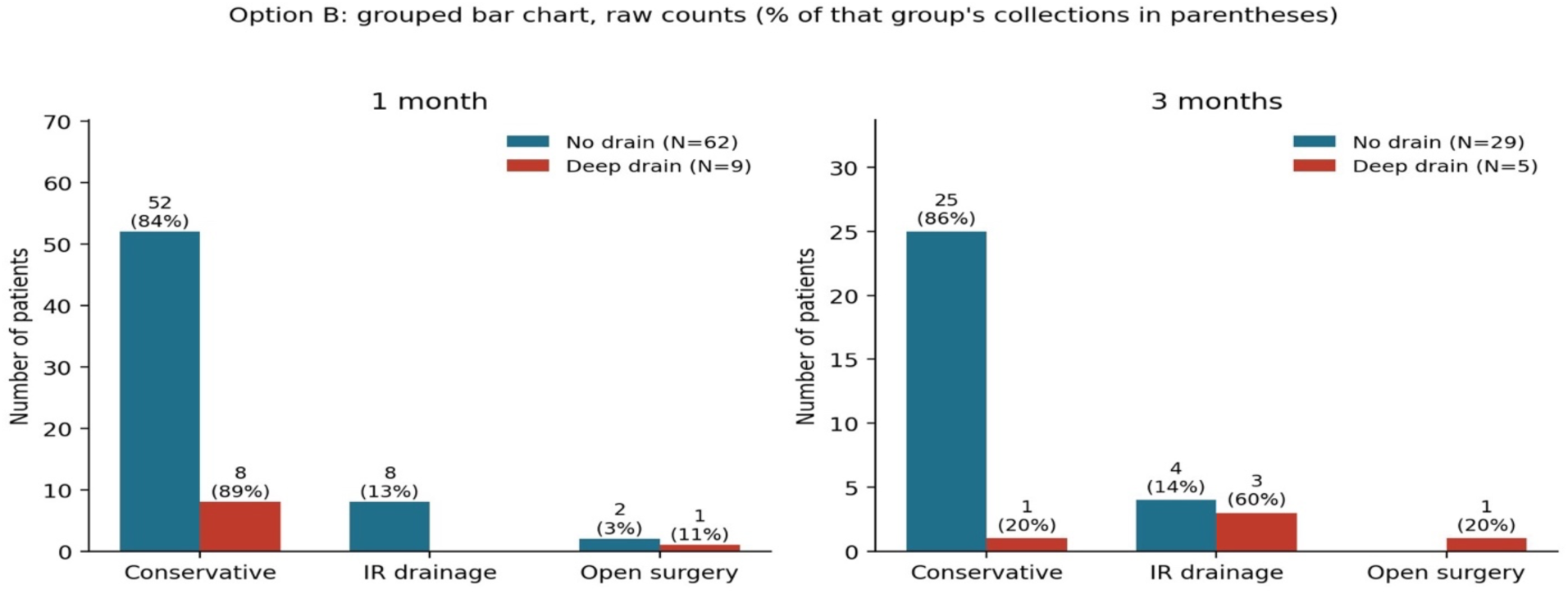
Management among patients with a peri-graft collection at 1 month (no drain n=62, deep drain n=9) and 3 months (no drain n=29, deep drain n=5).

### Interpretation of the deep-drain subgroup

The deep-drain subgroup had a prevalence trajectory similar in shape to that of the no-drain group. However, recipients selected for deep drainage had significantly higher BMI and, by definition, perioperative anticoagulation, and only five deep-drain recipients had a collection at 3 months.

The statistically significant difference in 3-month management should therefore be interpreted as an observation in a high-risk, clinically selected subgroup. It cannot establish that deep drains increased the need for intervention or that they were ineffective.

### Persistence and resolution

Individual-patient persistence and resolution could not be determined from the available aggregate data. A collection observed at 3 months may have been persistent from 1 month, newly developed after a negative 1-month study, or unrelated to an earlier collection. No individual-level incidence, persistence, or resolution rate was calculated (Figure 4). The fall in cross-sectional prevalence from 48.6% at 1 month to 23.3% at 3 months is not the same as 37 patients resolving: some of the 34 patients with a 3-month collection may be new, and some of the apparent net resolution may be masked by other new collections appearing.

**Figure 4.**
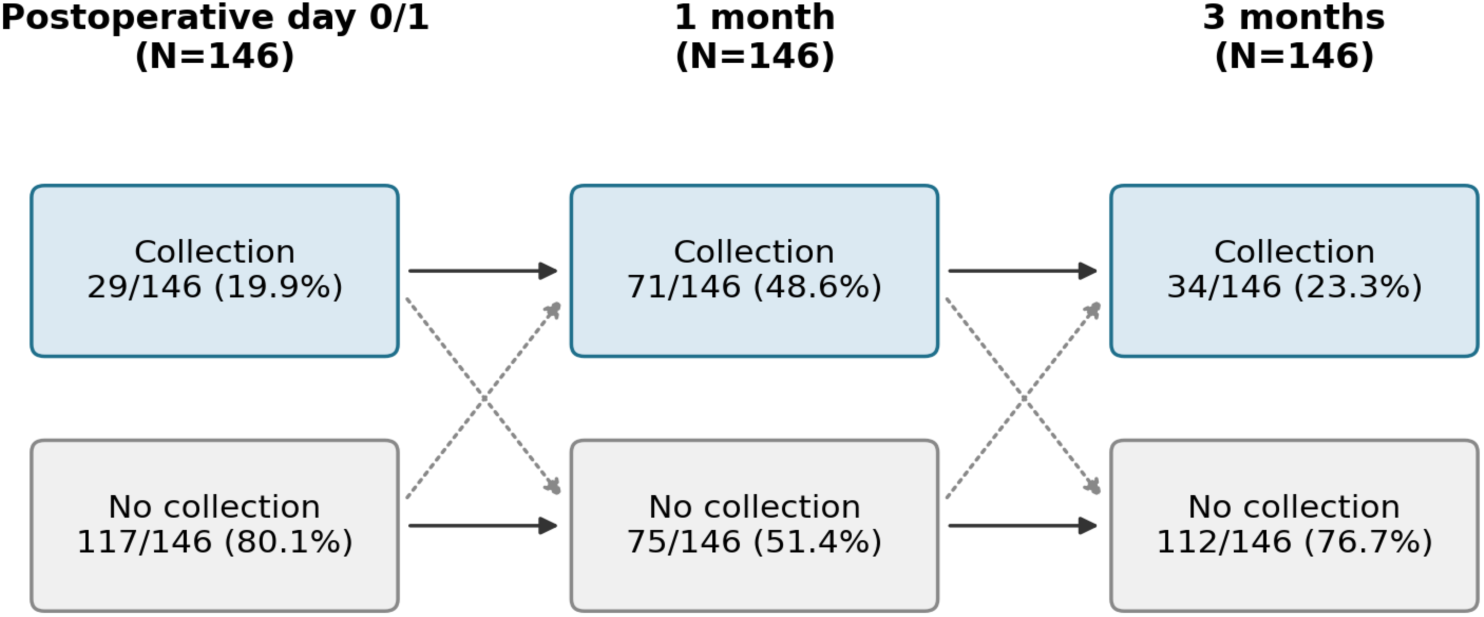
Bounds on collection transitions between imaging visits. Solid arrows connect observed cross-sectional counts; dotted arrows indicate transitions that cannot be excluded from aggregate data alone. Because only 29 patients had a collection on day 0/1, at least 42 of the 71 patients with a 1-month collection did not have one on day 0/1. Patient-level linkage between visits was not available, so persistence, resolution, and new formation in individual patients cannot be read from these counts. A patient-level alluvial diagram requires a linked dataset.

## Discussion

### Principal findings

In this serial-imaging cohort, peri-graft collections were common and time-dependent: about one in five recipients had one on postoperative day 0/1, one in two at 1 month, and one in four at 3 months.

For patients managed without a drain, most collections were handled conservatively at 1 month (83.9%) and 3 months (86.2%). Image-guided drainage was necessary in roughly 13% of cases at both intervals, and open surgery was rare. While these findings favor conservative management as a first-line approach for stable collections, conclusions are tempered by the absence of granular data regarding specific collection dimensions and clinical symptoms.

The deep-drain subgroup had significantly higher BMI than the no-drain group, and although 3-month invasive management was more frequent among deep-drain recipients with a collection, this was an exploratory association in a very small, indication-enriched subgroup and cannot be interpreted as a causal effect of drain placement.

### Comparison with previous work

Our 1-month collection prevalence is high compared with symptom-triggered series, which is expected when utilizing scheduled imaging rather than evaluating only symptomatic patients. For instance, in a large cohort undergoing routine graft sonography, collections were identified in 35.2% of recipients—with 8.3% requiring intervention—while neither the collections nor the interventions compromised long-term graft survival. Among our no-drain recipients, 16.1% (10 of 62) with a 1-month collection underwent invasive management (interventional radiology plus surgery), although direct comparisons remain challenging due to institutional differences in imaging schedules, definitions, and follow-up protocols.

On drain strategy, a 2024 meta-analysis of four retrospective cohorts (2,002 patients) found no significant reduction in reintervention (OR 0.59, 95% CI 0.16-2.23) or perinephric collections (OR 0.55, 95% CI 0.13-2.37) with prophylactic drainage and recommended a case-by-case approach ^11^. An earlier systematic review and several single-center studies agree ^8–10, 13^. However, one cohort could not be pooled because it reported only an adjusted odds ratio, and it found fewer peri-graft collections with drains after adjusting for BMI and other factors (OR 0.62, 95% CI 0.43-0.88) ^11^, and another series reported fewer lymphoceles and wound complications ^12^. Because our deep-drain patients were also selected by clinical risk, the evidence does not uniformly favor avoiding drains.

Early collections may be seroma, hematoma, or urinoma, while symptomatic lymphoceles usually appear between 2 weeks and 6 months ^4^. We therefore regard the 1-and 3-month time points as the informative ones. Our findings suggest that most screen-detected collections are benign and can be managed conservatively, and they add serial, protocolized imaging data under a predominantly no-drain protocol.

### Interpretation of the prevalence trajectory

The 1-month peak likely reflects scheduled imaging that detects collections developing during the early postoperative weeks. It should not be interpreted as evidence that collections identified on postoperative day 0/1 necessarily enlarged or persisted. The lower prevalence at 3 months is consistent with a reduction in the number of patients with a detectable collection at that visit; however, because patient-level imaging linkage was unavailable, the data do not establish which collections resolved, which persisted, and which developed de novo (Figure 4).

### Clinical meaning of the deep-drain subgroup

The deep-drain recipients represent a clinically meaningful group rather than a conventional treatment arm. Deep drains were used selectively in recipients with perioperative anticoagulation and other perceived risk factors, and the subgroup also had higher BMI, providing an additional axis of confounding.

The absence of a significant difference in collection prevalence at any time point suggests that the observed collection burden was not clearly different between groups, but the analysis is underpowered and cannot assess whether deep drainage prevented collections, modified their size or severity, or influenced symptoms.

The higher frequency of invasive management at 3 months may reflect the underlying risk profile of the subgroup, differences in collection characteristics, clinician thresholds for intervention, or chance, and should not be interpreted as drain-associated harm.

Differences in surgical technique, drain type, patient selection, anticoagulation regimen, imaging schedule, and definitions of lymphatic complications likely explain some of the variation across the studies discussed above^1–3,16,17^

### Clinical implications

Scheduled imaging identifies many collections that do not require invasive treatment. A small or asymptomatic collection identified at 1 month may reasonably be observed when there is no graft dysfunction, hydronephrosis, venous compression, infection, or clinically significant symptom^3–5^.

A collection present at 3 months should prompt reassessment of symptoms, size, anatomy, graft function, and fluid characteristics when sampling is performed, rather than automatic drainage.

The deep-drain subgroup highlights the need for prospective studies specifically targeting high-risk recipients; such studies should record the indication for drainage, anticoagulation regimen, BMI, collection size, symptoms, fluid type, graft function, and intervention thresholds.

### Strengths and limitations

Strengths include consecutive cohort inclusion, scheduled imaging at prespecified time points, explicit separation of prevalence from incidence, transparent denominators, and acknowledgment of confounding by indication.

### Limitations are as follows

First, the design is retrospective and single canter, with one surgical program and one imaging program. Second, the deep-drain group is small (n=22), non-randomly assigned and defined by anticoagulation and BMI, so drain effects cannot be estimated.

Third, imaging occurred at three time points, so resolution timing is interval-censored and new collections at 3 months cannot be distinguished from persistent ones without patient-level linkage. Fourth, follow-up ended at 3 months, and symptomatic lymphoceles can appear up to 6 months after transplantation^4^. Fifth, the imaging modality varied between ultrasound and CT, and day 0/1 screening detects small clinically irrelevant fluid.

Sixth, collections were not uniformly aspirated, so type may be misclassified. Seven, this analysis used only aggregate group-level data; standard deviations, standardized mean differences, adjusted odds ratios, and a patient-level graft-function comparison could not be produced and are deferred to a planned patient-level analysis of the same cohort.

## Conclusion

Peri-graft collection prevalence after kidney transplantation was highest at 1 month and lower at 3 months in a predominantly no-drain cohort. Most collections were managed conservatively, and invasive treatment was uncommon in the no-drain group. A clinically selected deep-drain subgroup had a similar prevalence pattern but higher BMI and a greater frequency of invasive management among the small number of recipients with a 3-month collection.

This comparison is non-randomized and exploratory and does not permit inference about the benefit or harm of deep drains.

They support a surveillance-first approach to asymptomatic collections and justify a prospective, protocolized study, including a randomized comparison of drainage strategies in anticoagulated recipients.

## Abbreviations

BMI: body mass index
CI: confidence interval
CT: computed tomography
IR: interventional radiology
POD: postoperative day
STROBE: Strengthening the Reporting of Observational Studies in Epidemiology

## Prior presentation

An abstract has been Submitted to ATC 2027

## Data availability

Data supporting the findings of this study are available from the corresponding author upon reasonable request.

## References

1. Golriz M, Klauss M, Zeier M, Mehrabi A. Prevention and management of lymphocele formation following kidney transplantation. Transplant Rev (Orlando). 2017;31(2):100–105. doi: 10.1016/j.trre.2016.11.001

2. Mihaljevic AL, Heger P, Abbasi Dezfouli S, Golriz M, Mehrabi A. Prophylaxis of lymphocele formation after kidney transplantation via peritoneal fenestration: a systematic review. Transpl Int. 2017;30(6):543–555. doi:10.1111/tri.12952

3. Mehrabi A, Kulu Y, Sabagh M, et al. Consensus on definition and severity grading of lymphatic complications after kidney transplantation. Br J Surg. 2020;107(7):801–811. doi:10.1002/bjs.11587

4. Richard HM 3rd. Perirenal transplant fluid collections. Semin Intervent Radiol. 2004;21(4):235–237. doi:10.1055/s-2004-861557

5. Lehner LJ, Hohberger A, Marschke L, et al. Analysis of risk factors and long-term outcomes in kidney transplant patients with identified lymphoceles. J Clin Med. 2020;9(9):2841. doi:10.3390/jcm9092841

6. Nguyen E, Minkovich M, Famure O, et al. Lymphoceles: impact on kidney transplant recipients, graft, and healthcare system. Can J Urol. 2021;28(5):10848–10857.

7. Eliminating Routine Drain Placement in Kidney Transplantation: Impact on Postoperative Collections and Outcomes. Davtyan H, Makki MM, Kumar K, Fernandez E, Kusakabe J, Pinna A, Refaai K. Am J Transplant. 2026;26(7 Suppl 1): S965. doi: 10.1016/j.ajt.2026.05.20838.

8. D’Souza K, Crowley SP, Hameed A, et al. Prophylactic wound drainage in renal transplantation: a systematic review. Transplant Direct. 2019;5(7):e468. doi:10.1097/TXD.0000000000000908

9. Sidebottom RC, Parsikia A, Chang PN, et al. No benefit when placing drains after kidney transplant: a complex statistical analysis. Exp Clin Transplant. 2014;12(2):106–112.

10. Farag A, Gaynor JJ, Serena G, Ciancio G. Evidence to support a drain-free strategy in kidney transplantation using a retrospective comparison of 500 consecutively transplanted cases at a single center. BMC Surg. 2021;21(1):74. doi:10.1186/s12893-021-01081-x

11. Cimen S, Guler S, Tennankore K, Imamoglu A, Alwayn I. Surgical drains do not decrease complication rates but are associated with a reduced need for imaging after kidney transplant surgery. Ann Transplant. 2016;21:216–221. doi:10.12659/aot.898260

12. Lakha AS, Ahmed S, Hunter J, O’Callaghan J. Prophylactic peri-nephric drain placement in renal transplant surgery: a systematic review and meta-analysis. Transpl Int. 2024;37:13030. doi:10.3389/ti.2024.13030

13. Vincenzi P, Gaynor JJ, Chen LJ, et al. No benefit of prophylactic surgical drainage in combined liver and kidney transplantation: our experience and review of the literature. Front Surg. 2021;8:690436. doi:10.3389/fsurg.2021.690436

14. Simforoosh N, Mohseni Rad H, Tabibi A, Gholamrezaie HR. Comparison between bipolar lymphatic vessels cautery and suture ligature in prevention of postrenal transplant lymphocele formation: a randomized controlled trial. Exp Clin Transplant. 2019;17(1):26–30.

15. El Hennawy HM, Safar O, Al Atta E, et al. Prophylactic sclerotherapy for lymphatic complications after living donor renal transplant: a single-center experience. Exp Clin Transplant. 2024;22(1):17–21.

16. Lundin C, Bersztel A, Wahlberg J, Wadstrom J. Low molecular weight heparin prophylaxis increases the incidence of lymphocele after kidney transplantation. Ups J Med Sci. 2002;107(1):9–15.

17. Sabagh MS, Mohammadi S, Ramouz A, et al. Validating consensus-defined severity grading of lymphatic complications after kidney transplant. J Clin Med. 2021;10(21):4858. doi:10.3390/jcm10214858

18. Wilson EB. Probable inference, the law of succession, and statistical inference. J Am Stat Assoc. 1927;22(158):209–212.

